# Iranian Dental Students and Specialists’ Knowledge and Attitude about Obstructive Sleep Apnea

**DOI:** 10.1101/2020.08.07.20170605

**Authors:** Shervin Shafiee, Ahmad Sofi-Mahmudi, Mohammad Behnaz, Hannaneh Safiaghdam, Soroush Sadr

**Affiliations:** Oral and Maxillofacial Surgery Resident, Shahid Beheshti University of Medical Sciences, Tehran, Iran; Students’ Research Office, Dental School, Shahid Beheshti University of Medical Sciences, Tehran, Iran; Assistant Professor, Orthodontics Department, Shahid Beheshti University of Medical Sciences, Tehran, Iran

**Keywords:** Obstructive sleep apnea, Knowledge, Attitude, Dentistry

## Abstract

**Introduction:** Obstructive sleep apnea is a relatively common sleep disorder, which leads to multiple sleep arousal and hypoxemia. It also has a significant socioeconomic impact. Dentists can have a role in screening as well as treating apnea by orthodontic devices. No study has evaluated the knowledge and attitude of dental health professionals about obstructive sleep apnea (OSA) in Iran. We aimed to measure knowledge and attitude among students and faculty members of Shahid Beheshti Dental School, Tehran, Iran about OSA.

**Materials and Methods:** We conducted a cross-sectional survey among residents and professors in oral and maxillofacial surgery, orthodontics, and oral medicine specialties and dental students. The Obstructive Sleep Apnea Knowledge and Attitude (OSAKA) questionnaire was used to obtain the information regarding knowledge and attitude. We used Chi-square, Kruskall Wallis, Mann-Whitney U test for statistical analysis. The data were analyzed by SPSS 22.0 and a p-value lower than 0.05 considered to be significant.

**Results:** One hundred ninety-seven participants, including 43 dental students, 68 dental residents, and 64 dental professors filled the survey. Mean knowledge score among all participants was 10.69±3.133. Overall, oral medicine and oral and maxillofacial surgery professors had significantly higher correct answer ratios in the knowledge section than fifth and sixth-year dental students (p<0.001). There was no significant difference among other groups (p>0.05). About attitude, 91% of respondents reported that OSA is an important or extremely important disorder. However, only 10.2% and 16.9% felt confident about the ability to manage patients with OSA and identifying patients at risk for OSA, respectively.

**Conclusion:** All of the participants had poor knowledge but a positive attitude towards OSA. This shows the necessity of better education about OSA.

## Introduction

Obstructive sleep apnea (OSA) is a relatively common sleep disorder affecting 17 to 22 percent of the population ^1^. It is characterized by multiple episodes of airway obstruction during sleep that leads to multiple arousals and hypoxemia ^2^. OSA is associated with hypertension, traffic accidents, and decreased quality of life^3^. In addition, it has a significant economic impact on society because of daytime sleepiness and works disability ^4^ More importantly, moderate to severe sleep apnea increases the risk of all-cause mortality ^2,5^. Five percent of Iranians were deemed high risk based on Berlin questionnaire criteria ^6^. However, it is considered to be an underdiagnosed disease as identifying patients is a challenge in public health^7^.

Dentists can be the first line of detecting and managing many diseases. Owing to frequent contact with people and the availability of simple questionnaire-based screening tools, dentists can play a significant role in screening OSA patients and referring them to sleep specialists ^8^. In addition, they may be involved in the treatment plan. The gold standard treatment for apnea is continuous positive airway pressure (CPAP) ^9^. It may have side effects and discomforts, and it may not be well tolerated. Dentists can contribute by preparing a mandibular advancement device for CPAP-noncompliant patients ^10^. Moreover, a variety of oral surgeries including uvulopalatopharyngoplasty (UPPP), hyoid advancement, mandible advancement, and etcetera can be performed by the maxillofacial surgeon on eligible patients ^11^.

Despite the importance of their role, dentists are reluctant to get involved in managing apnea patients probably because of fear of malpractice ^8^. There is a lack of adequate training in dental schools in both predoctoral and postdoctoral curricula^12^. Mean total predoctoral sleep curriculum time is 2.96 h in 49 USA dental schools^13^. This number diminishes to 1.2h in certain middle eastern countries^14^. In Iran, there is no specific the training in predoctoral and postdoctoral curriculum, and it is only briefly explained.

Therefore, we aimed to measure knowledge and attitude among undergraduate students and orthodontics, maxillofacial surgery and oral medicine residents and attending professors of Shahid Beheshti Dental School to provide an overview.

## Materials and Methods

We surveyed Shahid Beheshti Dental School (Tehran, Iran) students, residents, and attending professors of oral medicine, maxillofacial surgery, and orthodontics departments in April 2016 and used extra online forms for achieving desirable sample size for residents and attending professors. We chose these departments’ professors and residents as they are in contact with OSA patients more than other departments and have only students who completed basic sciences courses and were in their fifth and sixth year of dentistry were enrolled in this study. The questions were selected from OSAKA questionnaire ^15^ that is designed and validated in the United States for measuring knowledge and attitude of physicians towards sleep apnea. To obtain Persian version of OSAKA questionnaire, we used cross-cultural translation method. In brief, two authors translated the questionnaire into Persian separately. A review committee consisting of orthodontists and English language experts evaluated these two versions and chose one of them as the provisional version. Then two bilingual investigators back translated this version to English. Neither of them had access to the original version of the questionnaire. A committee compared these versions with the original version and assessed its equivalency and consistency to produce a pre-final version. We distributed the questionnaires among 25 respondents and then interviewed them to assess the understandability and accessibility of questionnaire. The final version of Persian translation was validated by the committee based on the assessment given by these 25 people.

The questionnaire consisted of two main sections, knowledge and attitude. Knowledge section had 18 questions with possible “true”, “false”, and “do not know” choices and covered subjects including epidemiology, pathophysiology, symptoms, diagnosis, and treatment of OSA. “Do not know” answers were considered as false. Attitude section had five questions. The first two questions were about the importance of the OSA in a five-level Likert item from “not important” to “extremely important” choices. The next three attitude questions were about confidency of identifying and managing the OSA patient in a five-level Likert item from “strongly disagree” to “strongly agree”.

The researchers distributed printed questionnaires among the targeted group in the dental school. The data was analyzed using SPSS 22 (SPSS, Inc., Chicago, IL). We used Chi-square test for comparing correct answers in different educational levels in each question. The total knowledge score was compared with Kruskal Wallis and Mann-Whitney U test. Then, the knowledge and attitude score were compared with Spearman correlation. We used a two-tailed test and α < 0.05.

## Results

### Characteristics

Of 242 questionnaires, 177 were handed out (response rate: 73.1%). Eighty-nine respondents were female (50.3 percent) and 88 were male (49.7) with a mean age of 32.44 years (SD=9.00) ranging from 21 to 55. The majority of respondents were dental residents (34.5 percent). Table 1 summarizes the demographic information of the participants. None of the participants, including OMF surgeons, had experience of apnea subspecialty training.

**Table 1.** Characteristics of participants

| Characteristics | Value |
| --- | --- |
| Age (Mean ± SD) | 32.44 ±<br>9.00 |
| Gender | N (%) |
| Male | 88 (49.7) |
| Female | 89 (50.3) |
| Degree | N (%) |
| Dental students | 43 (24.3) |
| Residents | 68 (38.4) |
| Oral and maxillofacial surgery | 22 (12.4) |
| **Orthodontics** | 22 (12.4) |
| **Oral and maxillofacial medicine** | 24 (13.6) |
| **Professors** | 66 (37.3) |
| **Oral and maxillofacial surgery** | 22 (12.4) |
| **Orthodontics** | 21 (11.9) |
| **Oral and maxillofacial medicine** | 23 (13) |

### Knowledge

Mean knowledge score among all participants was 10.69 ± 3.133, ranging from 8.43 ± 3.55 for fifth-year dental students to 12.39 ± 1.75 for OMF medicine professors. Overall, professors obtained a higher score compared to the students on average. The internal consistency of items on the knowledge scale was acceptable, with a Cronbach α of 0.64. The detailed information about knowledge questions is illustrated in Table 2.

**Table 2.**
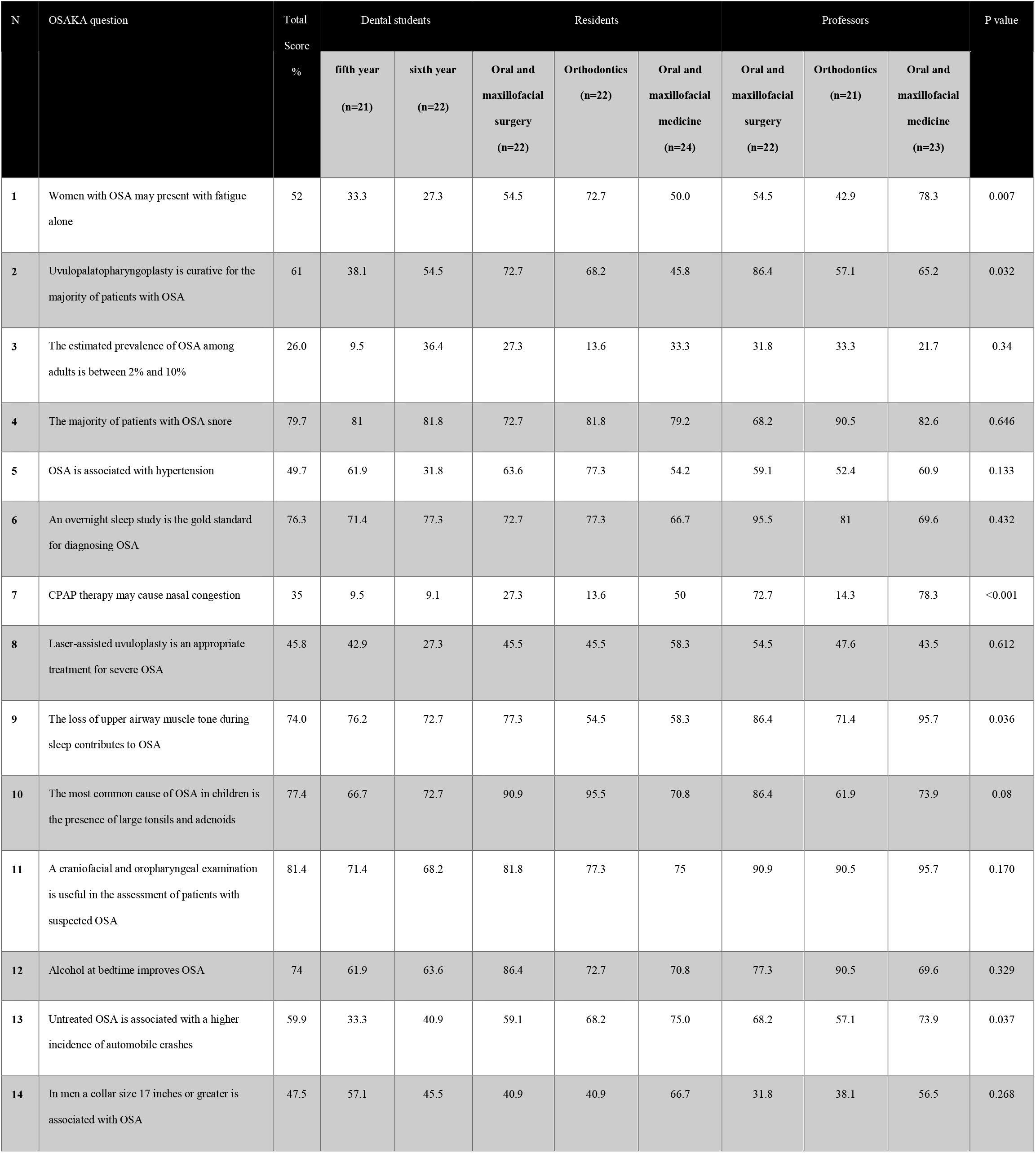
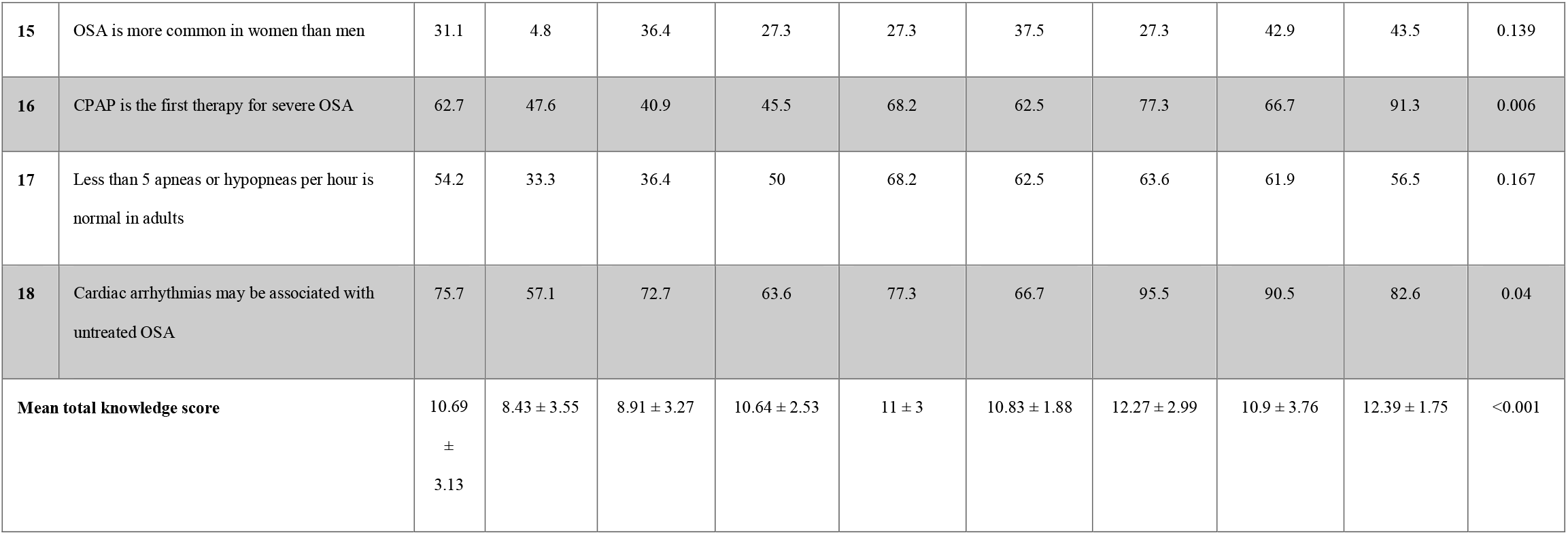
Knowledge questions by each type of respondents

More than two-thirds of respondents did not answer the questions about the prevalence of OSA and whether OSA is more common in women than men correctly (the highest correct answer ratio was among dental students in the sixth year and OMF medicine professors, respectively). Less than 10 percent of dental students answered the questions about whether CPAP therapy causes nasal congestion correctly.

About 80% of respondent answered the question about snoring of patients with OSA and usefulness of a craniofacial and oropharyngeal examination correctly (with the highest mean correct answer ratio of 90.5% and 95.7% among orthodontics professors and OMF medicine professors, respectively).

Seven knowledge items significantly differed among respondents, which were questions 1, 2, 7, 9, 13, 16, and 18. Of these items, question 7 that was about whether CPAP therapy may cause nasal congestion had the most significant difference (p < 0.001). In this item, OMF medicine professors mean correct answer ratio was significantly higher than all other respondents (p < 0.001). Overall, OMF medicine professors and OMF surgery professors had significantly higher correct answer ratios than fifth and sixth-year dental students (p < 0.001). There was no difference in total knowledge score with regard to gender (p=0.941).

### Attitude

Ninety-one percent of respondents reported that OSA is an important or extremely important disorder, and 88.7% answered that identifying patients with possible OSA is important or extremely important. In questions about confidence, only 10.2% and 16.9% felt confident about the ability to manage patients with OSA and identifying patients at risk for OSA, respectively. Whereas, 34.5% of respondents felt confident in their ability to manage patients of CPAP therapy. Total attitude score was higher in male responders (p<0.001). The correlations between all attitude items, total attitude score and total knowledge score were significant (Table 3).

**Table 3.**
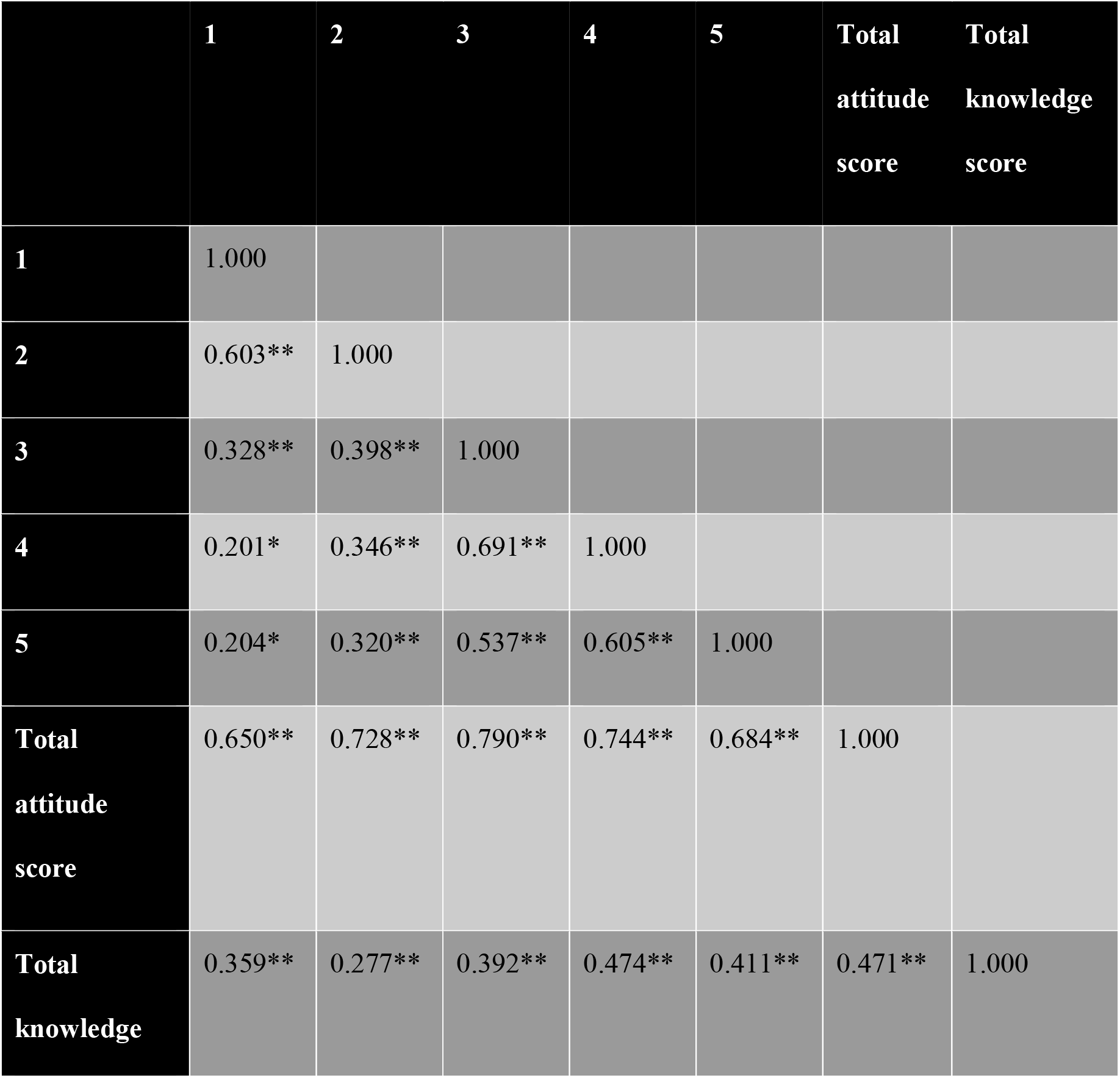
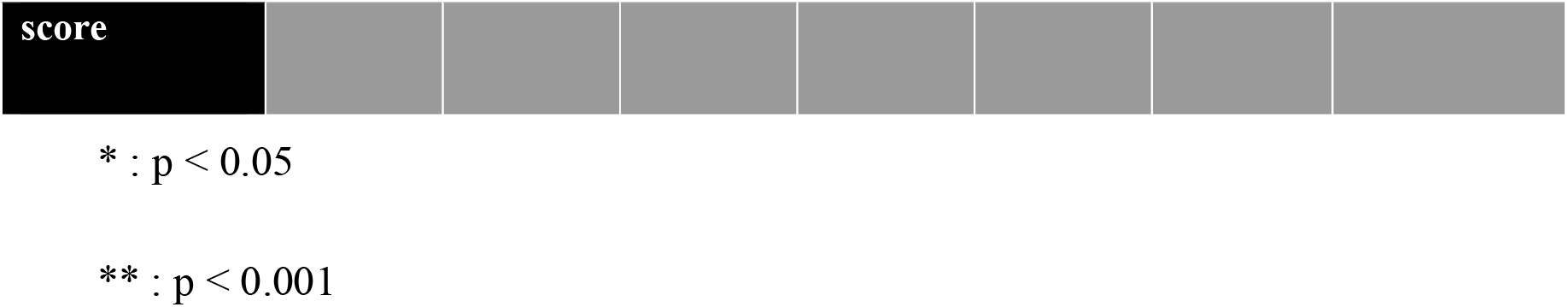
Correlation among attitude items and total attitude and knowledge score

## Discussion

OSA is a highly underdiagnosed condition that may lead to multiple health problems. ^16, 17^ Despite noticeable prevalence shown in different studies in Iran, this disorder still remains under-recognized by different specialties, among them dentists. ^18, 19^ As dental professionals are determined to treat patients comprehensively, they should be concerned about OSA, with high blood pressure, depression, impaired quality of life and increased mortality among its consequences. ^20^ Moreover, certain associated risk factors such as edentulism and possible treatments such as mandibular advancement are directly linked to a dental practice, thus the clinical relevance of OSA.^21,22^ While no studies looked at how frequently dentists were involved in diagnosing OSA, available data suggest that there is a lack in referral and management of these patients. Therefore, in this study, we sought to evaluate the knowledge and attitude of Iranian dental students, resident, and attendees towards OSA to reveal one of the possible reasons behind this underdiagnosis.

Obtained results show that mean total knowledge score was far from optimal. Our results fall below those of the study by Talaat et al. that evaluated fifth-year dental students’ knowledge of sleep medicine in the second Sharjah International Dental Student Conference in April 2014.

Nearly 29% of the respondents were in the high score group, whereas 70.8% scored low in knowledge of sleep-related breathing disorders. ^14^ In another survey, 40% of the participants, who were general dentists in the US, reported that they knew little or nothing about oral appliance treatments for OSA patients. ^12^ In another study in Finland, Vuorjoki-Ranta et al. argued that the risk factors, signs and symptoms, and consequences of OSA were overall well recognized among dental professionals regardless of the years of practice. For example, 41.8% of the participants, including GPs and specialists identified hypertension as a consequence of OSA by choosing “totally agree”, while in our study, 49.7% admitted there was a correlation. ^23^ However, the results can be compared because of the different designs of the used questionnaires. Similar to the original OSAKA survey, in this study, there was no relation between gender and knowledge. ^15^ Respondents scored the least on epidemiological questions such as the prevalence of OSA or its gender predilection. However, nearly 80% of them, considered a craniofacial and oropharyngeal examination useful in the assessment of patients with suspected OSA. Although a trend of increased knowledge was observed with increased years of education, no significant difference was observed except that OMF medicine professors and OMF surgery professors had significantly higher correct answer ratios than fifth and sixth-year dental students.

Despite an insufficient level of knowledge, Ninety-one percent of respondents reported that OSA is an important or extremely important disorder, and the majority considered that identifying patients with possible OSA is important or extremely important. This finding is of particular importance as it can be concluded that dentists are willing to intervene in the management of OSA patients if they are given required training. It has been argued that the reluctance of dental professionals to get involved in the diagnosis and treatment of OSA patients could be attributed to their low level of confidence. ^24^ Only about a quarter of participants felt confident about their ability to manage patients with OSA and identifying patients at risk for OSA, according to our results. A significant correlation was also observed between all attitude items, total attitude score and total knowledge score. In other words, the more dentists know about the condition and its risk factors and consequences, the more willing they are to help. This finding is supported by several similar surveys using OSAKA questionnaire.^15,24-27^

The low level of knowledge regarding OSA as demonstrated in this study is alarming especially considering the complications of OSA. Due to an increase in the associated risk factors such as obesity, more emphasis should be put on training dentists who will appropriately identify OSA patients and actively participate in their referral or treatment. ^28^ The average educational time spent on sleep medicine in US dental schools is reported to be 3.92 hours for pre-doctoral dental program and 1.55 for dental hygienist program. ^13, 29^ Similarly, Japanese pre-doctoral dental curriculum consists of 3.8 hours of training on sleep medicine.^30^ A lower educational time is devoted to the subject in middle east dental school, averaging about 1.2 hours. ^14^ This unsatisfying instruction time drops to null in dental curricula of undergraduate and graduate programs of Iran dental schools, although there is not yet a national survey conducted. The under-representation of the importance of sleep medicine and OSA is a plausible reason for this insufficient emphasis of curricula on the subject. Therefore, our study sheds light on the lack of knowledge among dental graduates and the importance of integrating sleep medicine into dental curriculum. Studies have explored various interventions to improve dentists’ knowledge and encourage them to treat OSA patients. For example, Tsuiki et al, proposed an oral appliance therapy as an educational activity at the University of British Columbia^31^ while Yamamoto et al. innovated a new seminar consisting of a 60-min didactic lecture and a 2-h instructional practical training for postgraduate resident education in Nagasaki University. ^32^ It has also been suggested that updating the medical history form in a way that allows screening for sleep-disorders by dental students to help identify OSA patients. ^33^ There should be courses available for graduated dentists to continue education on sleep medicine and OSA, as suggested by Canadian dental sleep medicine professionals. ^34^ The authors also suggest the inclusion of information on OSA in national dental textbooks. Teodorescu et al. found that sleep medicine coverage in medical textbooks was less than 2% and had not been increased since the 1990s. ^35^ A similar study on dental textbooks is also prompted.

## Conclusion

In conclusion, senior dental students, postgraduate residents and attending professors of OMF medicine, OMF surgery and orthodontics showed poor knowledge but a positive attitude towards obstructive sleep apnea. This finding points to the need for better education in the field of sleep medicine in dental schools of Iran. With a response rate of 81.4%, this study conducted in one of the most prominent dental schools of Iran could help clarify the overall condition. However, the results obtained here are not to be generalized and are better be verified by other studies. Further studies should also focus on evaluating the knowledge of other specialties involved in treating OSA patients, looking at specific interventions to improve knowledge.

## Data Availability

Data will be available upon request.

